# Quantitative liver health imaging impacts surgical decision making and improves clinical outcomes in colorectal liver metastasis surgery

**DOI:** 10.1101/2023.04.21.23288453

**Authors:** Fenella Welsh, Pulkit Sethi, Senthil Sundaravadnan, Ben Cresswell, John Connell, Sina Knapp, J Michael Brady, Rajarshi Banerjee, Myrddin Rees

## Abstract

**Introduction:** Post-hepatectomy liver failure (PHLF), driven by insufficient volume and quality of the remnant liver following an operation, is a significant clinical problem that is currently underserved by pre-operative assessment methods. Clinical management and a patient’s recovery from post-operative liver related complications results in a protracted stay in hospital.

**Methods:** 91 patients with colorectal liver metastasis being considered for liver resection were recruited onto the Precision1 trial. The imaging report from an additional non-quantitative multiparametric MRI (mpMRI) scan was examined and used to alter surgical decision making. Patient outcomes were monitored and evaluated against a standard of care comparator dataset blinded to mpMRI scan results.

**Results:** Previously undiagnosed liver disease activity or elevated liver fat was detected using mpMRI in 23% of patients, whereas the liver health was unexpectedly good in 7% of patients; this resulted in a change to surgical plan in 29% of cases. The incidence of protracted (over 14 days) length of stay was reduced from 5% to 1% following the introduction of mpMRI reports into surgical decision-making process.

**Conclusion:** mpMRI is a safe method to evaluate liver health in patients being considered for liver resection. Surgical decision making can be altered to achieve a safer treatment strategy resulting in shorter hospital stays for patients.

## Introduction

The incidence of liver tumours is on the rise, driven by obesity, non-alcoholic steatohepatitis, viral hepatitis, and metastases from colorectal cancers, representing a substantial global health care challenge^1,2^. Among the different types of treatment options, liver resection is the major curative therapy for patients with primary liver cancer^3^ and for those with liver metastases^4^. However, despite improvements in surgical techniques, surgery for colorectal liver metastases is still considered a high-risk procedure owing to perioperative morbidity^5^; up to 26% of patients may suffer from surgery-related complications^6^. Post-hepatectomy liver failure (PHLF), driven by insufficient volume and quality of the remnant liver following an operation, is a critical clinical problem that is currently underserved by pre-operative assessment methods. Clinical management and a patient’s recovery from PHLF results in a protracted stay in hospital, often surpassing the median length of stay of 7 days in UK^7^.

Despite the high rates of curative outcomes, pre-surgery assessment criteria result in fewer than 25% of patients with primary or secondary liver cancer being considered eligible for surgery. Along with factors such as number and stage of tumours, the patient’s physical status and the assumed function of the liver (Child-Pugh score) are often involved in the decision making to rule out a candidate as not suitable for liver surgery as a first line treatment^8^ and to assess likelihood of post-surgery complications. Predicted liver performance after hepatectomy is dependent on the function and regenerative capacity of the remaining tissue (future liver remnant volume, FLR). Accurate assessment of these factors is therefore essential to ensure suitable patients are recognised and can access the most relevant treatment options. Additionally, accurate understanding of liver health may prevent surgical candidates from undergoing alternative, lengthy and costly treatments, and unsuitable patients being placed at greater risk for poor outcomes. There is therefore an unmet need to more accurately characterise the suitability of an individual patient by using a more personalised and objective measure of their current liver health.

Non-invasive assessment of liver health using quantitative imaging is a technique becoming widely adopted across hepatology and gastroenterology. Liver fat and liver disease activity can be measured using quantitative imaging methods (proton density fat fraction (PDFF), and iron corrected T1 mapping (cT1)) as radiological features of liver state and of disease and are associated with standard invasive markers of pathology from biopsy samples. These non-invasive, quantitative multiparametric MRI (mpMRI) metrics are now recommended in clinical guidelines for the assessment of fatty liver disease and cirrhosis^9–11^. In the assessment of liver health and function, cT1 may be of particular relevance given it has been associated with liver related clinical outcomes^12^ and cardiovascular related outcomes^13^. We have previously demonstrated in an observational study that cT1 and the future liver remnant both independently and combined predicted the length of stay of patients following liver resection^14^.

The aim of this study was to assess how the availability of information describing liver fat and liver disease activity would affect the clinical decision-making in patients being considered for liver resection. By providing liver health assessment via mpMRI, this interventional study aimed to optimise personalised healthcare for liver cancer patients^15^. We hypothesise that mpMRI of the liver offers better information as to the risk of peri-operative complications and will impact clinical decision making to avoid protracted hospital stays for patients undergoing liver surgery.

## Methods

### Study design

A prospective observational cohort study in patients with colorectal liver metastases who were referred for liver resection. The Precision1 study^15^ (https://clinicaltrials.gov/ct2/show/NCT04597710) was approved by the Brighton and Sussex Research Ethics Committee (REC Reference 20/PR/0222). All clinical investigations were conducted in accordance with the Declaration of Helsinki 2013.

### Patient eligibility criteria

Patients aged 18 and over awaiting surgery for liver metastasis were eligible for enrolment. 111 participants (Figure 1, Consort diagram) were assessed for eligibility in the trial at a tertiary referral liver surgery centre: Hampshire Hospitals NHS Foundation Trust, Basingstoke, UK. 91 participants were enrolled with informed consent, 20 did not enrol due to not fulfilling eligibility criteria. All 91 participants underwent a mpMRI scan prior to the multidisciplinary team (MDT) meeting. A surgical plan was established and recorded during the MDT prior to seeing the mpMRI reports and then reviewed after seeing the reports. Changes in the surgical plan were recorded. 11 participants underwent a two-stage treatment with an additional mpMRI scan before the second intervention.

**Figure 1:**
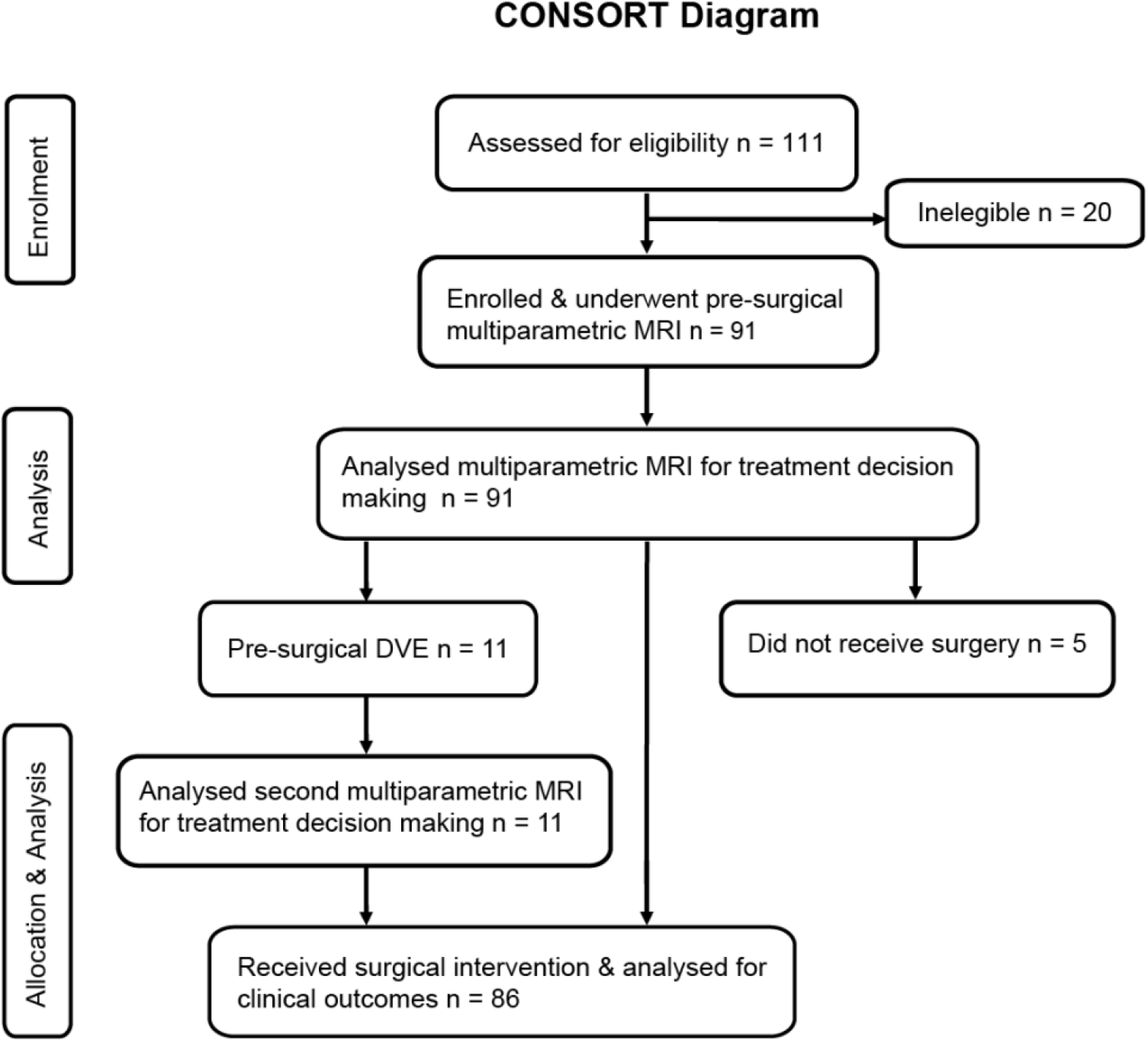
CONSORT Diagram of Precision1 patients from recruitment to allocation and analysis.

### Data collection

Data were collected according to the Precision1 protocol (Welsh et al., 2022)^15^. Data included limited medical history (regarding previous cancer diagnosis, liver diseases, comorbidities, recent chemotherapy regimens, alcohol intake, and smoking); blood test information (biochemistry, renal profile, liver function test, clotting (INR), haematology, tumour markers); pathological reports of tumour explants; mpMRI results (liver disease activity (cT1), liver fat (PDFF)). All participants underwent a 15-minute abdominal multiparametric MRI examination with the Hepatica image acquisition protocol. Liver disease activity was assessed with iron corrected T1 mapping^16^, with a reference value of 800ms being considered as the upper limit of normal^17^ (Andersson et al, 2021), liver fat measured using proton density fat fraction (PDFF) with a value of 5% considered the upper limit of normal. Patient questionnaires were obtained prior to surgery (utility of mpMRI report for disease understanding and expected length of stay in hospital); and 12 months after surgery (recovery, tumour recurrence).

### Comparison with historical surgical outcomes

A standard of care comparator dataset was curated from a clinical trial at the same centre during 2017-2018 involving a similar patient cohort (Table 1) with surgical resection for secondary metastatic and primary liver tumours at the same tertiary referral centre with the same operators. mpMRI scans were collected for this cohort but the clinical team were blinded to these results.

**Table 1:** Demographics and clinical characteristics of participants in Precision1 trial and a standard of care comparator. Median (IQR). Aspartate transaminase (AST); alanine aminotransferase (ALT); alkaline phosphatase (ALP); gamma-glutamyl transferase (GGT); alpha fetoprotein (AFP).

|  | Precision1 | Comparator | P value |
| --- | --- | --- | --- |
| Number of participants | 91 | 97 |  |
| Age | 66 (57-73) | 63 (56-72) | 0.795 |
| Sex male:female | 57:34 | 67:30 | 0.437 |
| BMI (kg/m<sup>2</sup>) | 26.9 (23.0-31.0) | 26.0 (23.4-29.1) | 0.312 |
| Pre-diagnosed liver disease | NAFLD (2) | Hepatitis C (1) |  |
| Tumour diagnosis | Colorectal liver metastasis (91) | Colorectal liver metastasis (91)<br>Liver metastasis (other) (3)<br>Hepatocellular carcinoma (2)<br>Cholangiocarcinoma (1) |  |
| Pre-operative chemotherapy | 85.7% | 19.6% |  |
| Pre-operative chemotherapy cycles | 6 (4-8) | 6 (4-13) |  |
| Baseline laboratory value<br>(normal reference values) |  |  |  |
| Platelets (10<sup>9</sup>/L) (150-400) | 236 (194-278) | 230 (191-292) | 0.794 |
| Bilirubin (μmol/L) 3–20 | 9 (7-13) | 9 (7-12) | 0.812 |
| Albumin (g/L) (33-49) | 38 (36-40) | 39 (37-41) | 0.163 |
| AST (U/L) (5-30) | 28 (23-34) | 26 (22-32) | 0.893 |
| ALT (U/L) (5-30) | 27 (21-35) | 24 (18-33) | 0.042 |
| ALP (U/L) (50-100) | 100 (85-134) | 90 (72-112) | ** 0.00481 |
| GGT (U/L) (6-50) | 43 (22-68) | 39 (26-69) | 0.271 |
| Prothrombin time (s) (12.5-14.4) | 13 (12-14) | 12 (12-13) | 0.153 |
| AFP (kU/L) (<10) | 2.6 (1.4-3.6) | 2.5 (1.5-4.7) | * 0.0439 |
| CA19-9 (kU/L) (0-33) | 18 (9-43) | 25 (7-55) | 0.184 |

### Statistical analysis

Descriptive statistics were used for data analysis. Results were expressed as mean with standard deviation (SD) for continuous normally distributed variables, as median and IQR for non-normally distributed data and as counts and/or percentages for categorical variables. The impact of mpMRI on the planned surgery was assessed using the percentage of cases changed between initial plan and review of mpMRI reports and explored visually using scatter and bar plots to assess how the mpMRI measurements led to change. The group-wise comparison in protracted length of hospital stay (≥14 days) between the Precision1 cohort and the comparator cohort was expressed as percentage of each cohort and was tested for significance using the Fisher’s exact test. For all tests, a p<0.05 indicated statistical significance. All statistical analysis was performed with R software (v 4.1.1; R Core Team 2021).

## Results

### Precision1 Participants

91 participants gave informed consent and enrolled onto the Precision1 study. 86 patients were deemed suitable for liver surgery and 5 were of too poor health to proceed with surgery; 11 of the 86 patients first underwent pre-surgical Dual Vein Embolisation (DVE) before receiving a second mpMRI for further treatment decision making. Clinical outcomes were analysed for all 86 patients who underwent surgery. The flow of participants through the study is depicted in Figure 1.

### Patient Characteristics

The demographics and clinical characteristics of the patient populations are shown in Table 1. For Precision1, 57 male and 34 female patients had colorectal liver metastases with a median age of 66 years (interquartile range, IQR, 57-73) and a median BMI of 27 kg/m^2^ (IQR 23–31). Only 2 of 91 had previously been diagnosed with chronic liver disease, whilst 89 of 91 participants were not known to have any diagnosed chronic liver disease.

The Comparator dataset consisted of 67 male and 30 female patients diagnosed with a median age of 63 years (IQR 56-72), and their median BMI was 26 kg/m^2^ (IQR 23-29). 91 patients had colorectal liver metastases, three with liver metastases other than from colorectal cancer, two with hepatocellular carcinoma, and one with cholangiocarcinoma. 1 patient presented with pre-diagnosed hepatitis C. The liver function tests and tumour markers in the Precision1 and Comparator patients are described in Table1, the ranges of operation types in each cohort are considered comparable.

### mpMRI liver health measurement

The average liver disease activity (cT1) and liver fat (%) was 752ms (±77ms) and 6.8% (±7.2%) respectively in the Precision1 cohort and 728ms (±67ms) and 7.2% (±7.0%) in the comparator cohort. 25.3% of Precision1 patients and 14.4% of patients of the Comparator cohort presented cT1 values above normal range (800ms). 35.2% of Precision1 patients and 35.1% (34 of 97 patients) of patients of the Comparator cohort presented PDFF values above normal range (5.6%) (see Table 2). The mpMRI results of the comparator group were blinded to the clinical team.

**Table 2:**
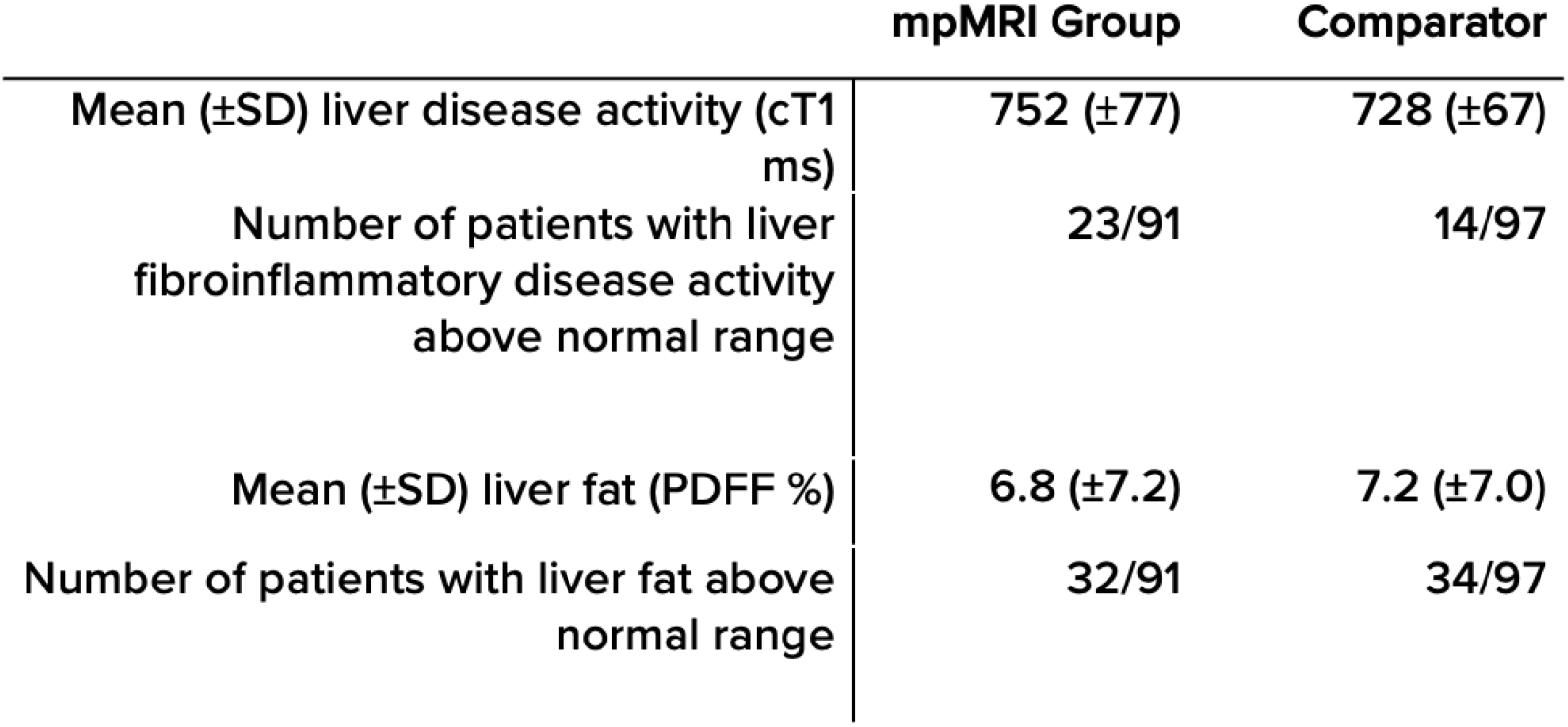
mpMRI liver health measurements of participants in Precision1 trial and a standard of care comparator. Mean±SD.

|  | mpMRI Group | Comparator |
| --- | --- | --- |
| Mean (±SD) liver disease activity (cT1 ms) | 752 ($\pm 77$) | 728 ($\pm 67$) |
| Number of patients with liver fibroinflammatory disease activity above normal range | 23/91 | 14/97 |
| Mean (±SD) liver fat (PDFF %) | 6.8 ($\pm 7.2$) | 7.2 ($\pm 7.0$) |
| Number of patients with liver fat above normal range | 32/91 | 34/97 |

#### Clinical utility

Examination of mpMRI reports resulted in a change of surgical plan in 29 cases (Figure 2), whereas after reviewing the remaining 73 reports, the liver health information increased confidence in the original surgical plans which were unchanged.

**Figure 2:**
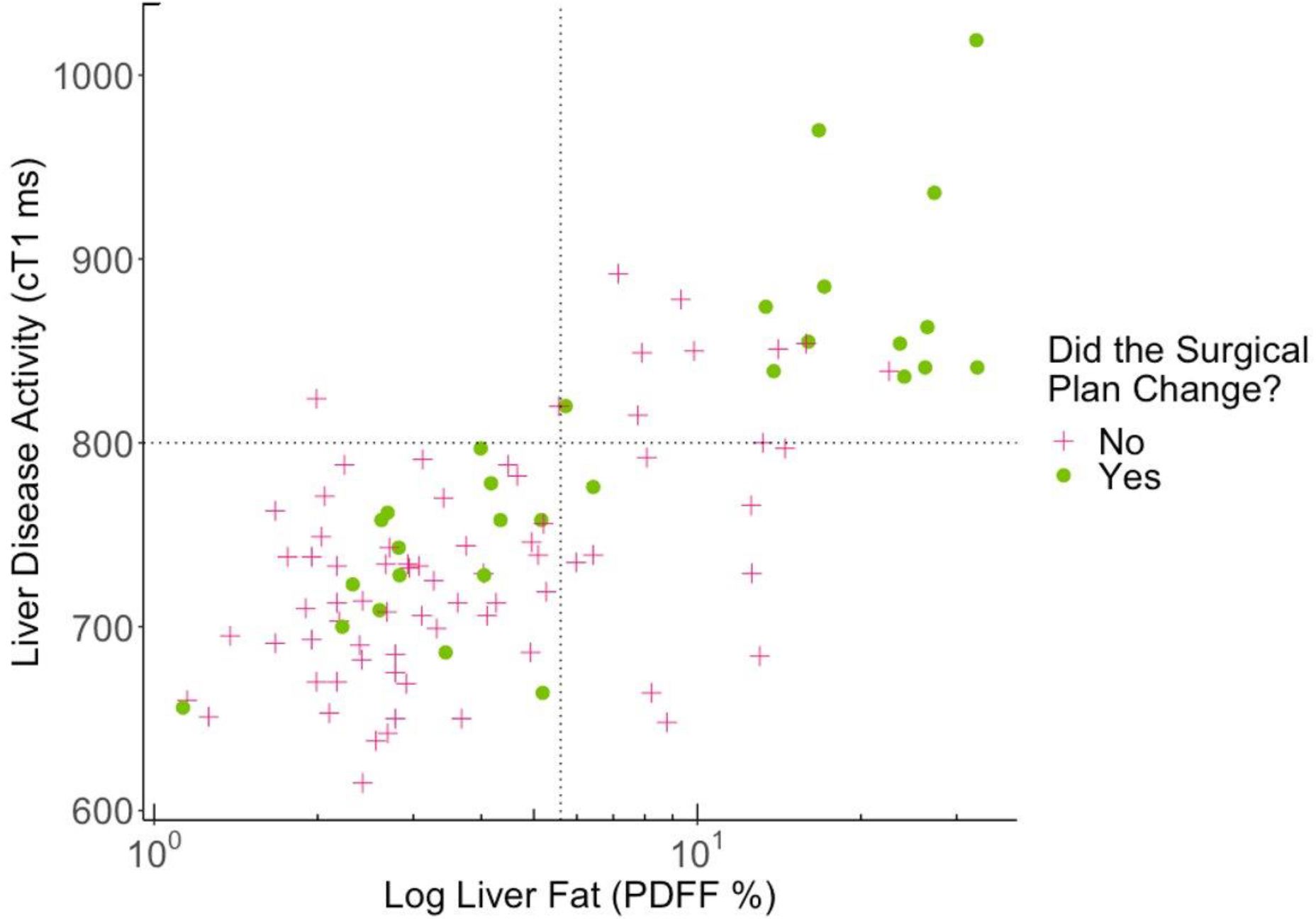
Changes in surgical plan from original plan (green dot) following examination of mpMRI report. Dotted lines indicate upper limit of normal for liver fat and liver disease activity biomarkers.

To understand better the characteristics of the cases with altered surgical procedures, cT1 and liver fat levels were analysed and categorised into four groups according to the reference range for cT1 (healthy range <800 ms) and liver fat (healthy range <5.6%): 68 patients were classified within both healthy ranges of these biomarkers; of these, the original surgical plan was confirmed in 53 cases, whereas 15 scans altered the surgical plan. 23 patients (22.5%) displayed both liver fat and cT1 values outside the healthy range, 13 of these patients had an altered the surgical procedure and review of 10 scans confirmed it. Of the 10 mpMRI scans (9.8%) with only elevated liver fat values, the surgical plan was changed in one case, whereas it was confirmed in 9 cases. One mpMRI scan classified as elevated cT1 values only and it confirmed the surgical plan (Figure 2). This is described in the four quadrants of Figure 2, defined by the upper limit of normal thresholds. The lower left corresponds to “good” liver state where surgical procedures were modified in these cases to be more extensive. The upper right corresponds to “poor” liver state - surgical procedures were modified in these cases to be more conservative. The co-incidence of elevated liver disease activity and liver fat changed the surgical plan more often than elevated liver fat alone.

Of the 29 patients who had a change in their surgical plan, eight had unexpectedly good liver health, resulting in a more extensive resection to improve the chances of curative resection; four of these patients had an initial surgical plan of a two stage approach. Five patients had a more conservative surgery owing to the perceived risk of a poor post-operative recovery period from liver insufficiency (Figure 3). The primary surgical plan was changed to a two-stage plan in 11 patients after reviewing their mpMRI scans to ensure the improved health of the remnant liver. Two scans indicated the change of the surgical plan in other ways (specific dietary changes prior to surgery) (Figure 3). Two illustrative cases are shown in Figure 4.

**Figure 3:**
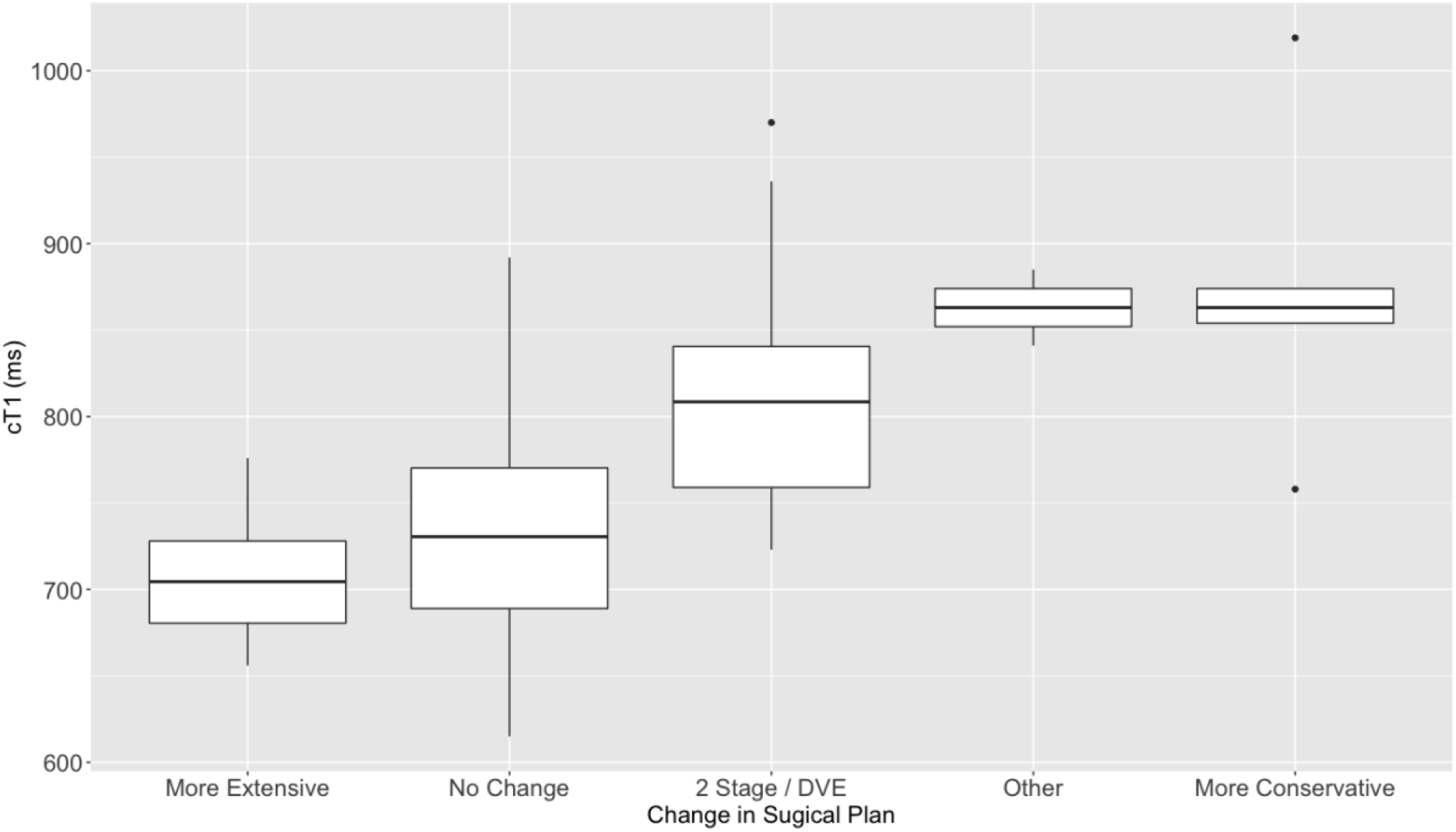
The change in surgical plan of 86 patients upon seeing liver health mpMRI report. Patients with poor liver health (cT1 above upper limit of normal (ULN)) underwent more conservative resections and 2 stage resections including dual vein embolisation (DVE). Eight patients with no signs of liver disease activity underwent more radical resection (green).

**Figure 4.**
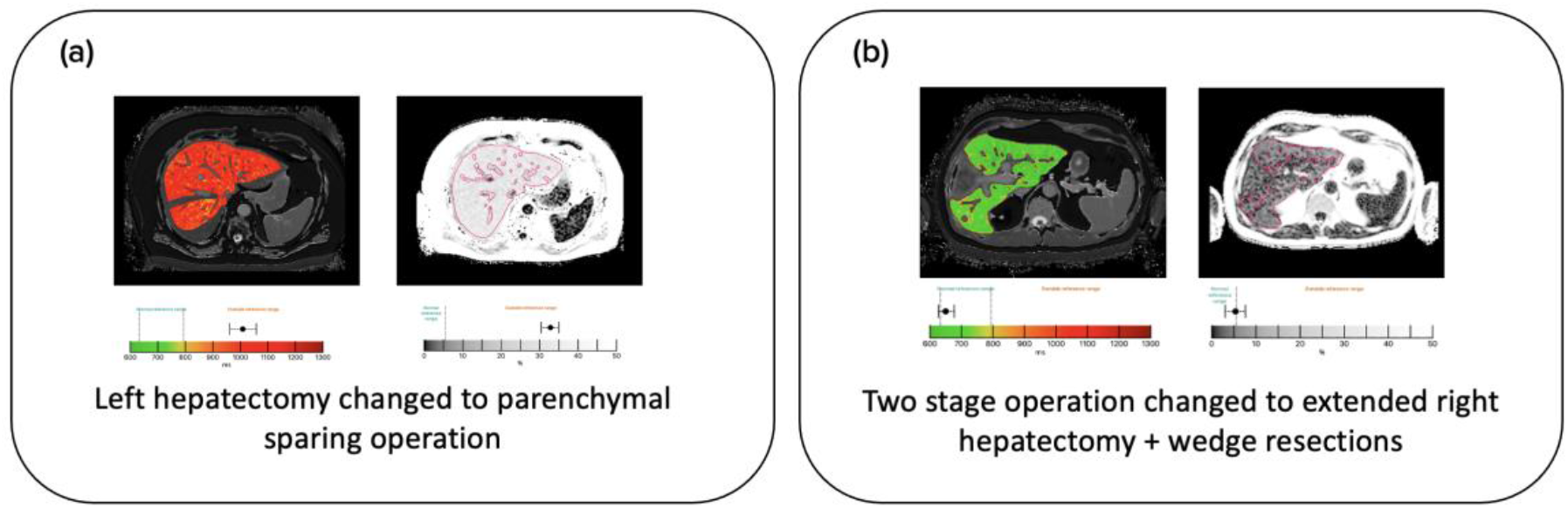
Example images showing change in surgical decision making after examining mpMRI report. (a) A patient presenting with liver disease activity and high liver fat underwent with parenchymal sparing operation preserving segment 2 and residual left lobe. (b) A patient presenting with an unexpectedly healthy liver underwent an extensive resection rather than a two stage operation.

**Figure 5.**
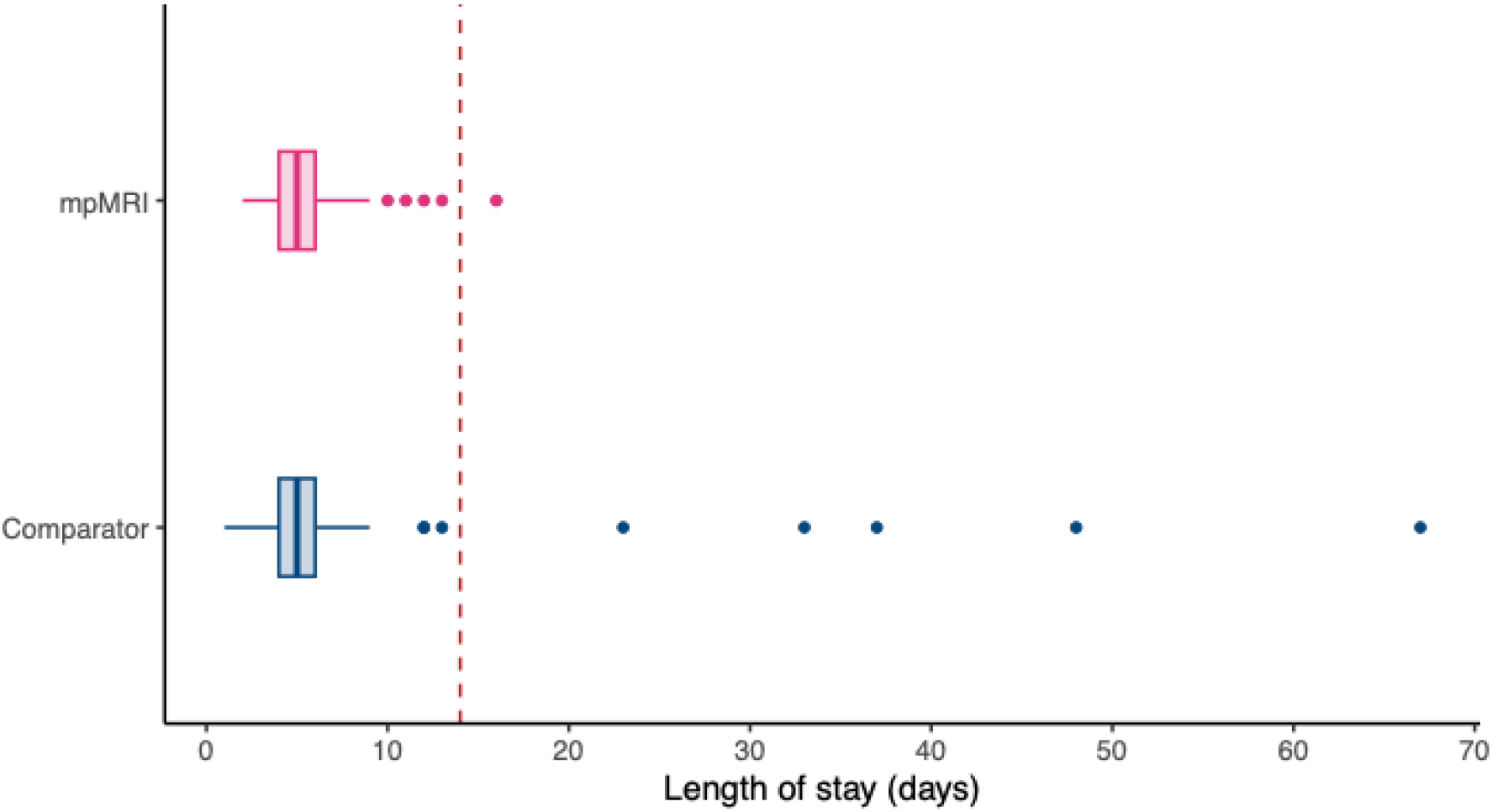
Length of stay in hospital after surgical resection for liver cancer. Five patients in the Comparator dataset required a protracted length of stay owing to post-operative complications.

## Clinical outcomes

### Length of stay

The mean (±SD) length of stay in the Comparator dataset was 6.7 days (±9.1); after the introduction of the mpMRI reports, the mean (±SD) length of stay was 5.3 days (±2.1) (p=0.147). Notably, the proportion of participants who had a protracted length of stay (>14 days) was considerably greater in the comparator dataset (5%) than the Precision1 cohort (1%) after the introduction of the quantitative imaging reports (P=0.136).

### Patient centred outcomes

Patients were asked about their understanding of diagnosis and treatment following provision of the mpMRI report. 89% of patients found the images helpful or very helpful in describing their diagnosis and treatment (Figure 6). 78% of patients confirmed that they would like to have a personal copy of the mpMRI report (Figure 6).

**Figure 6.**
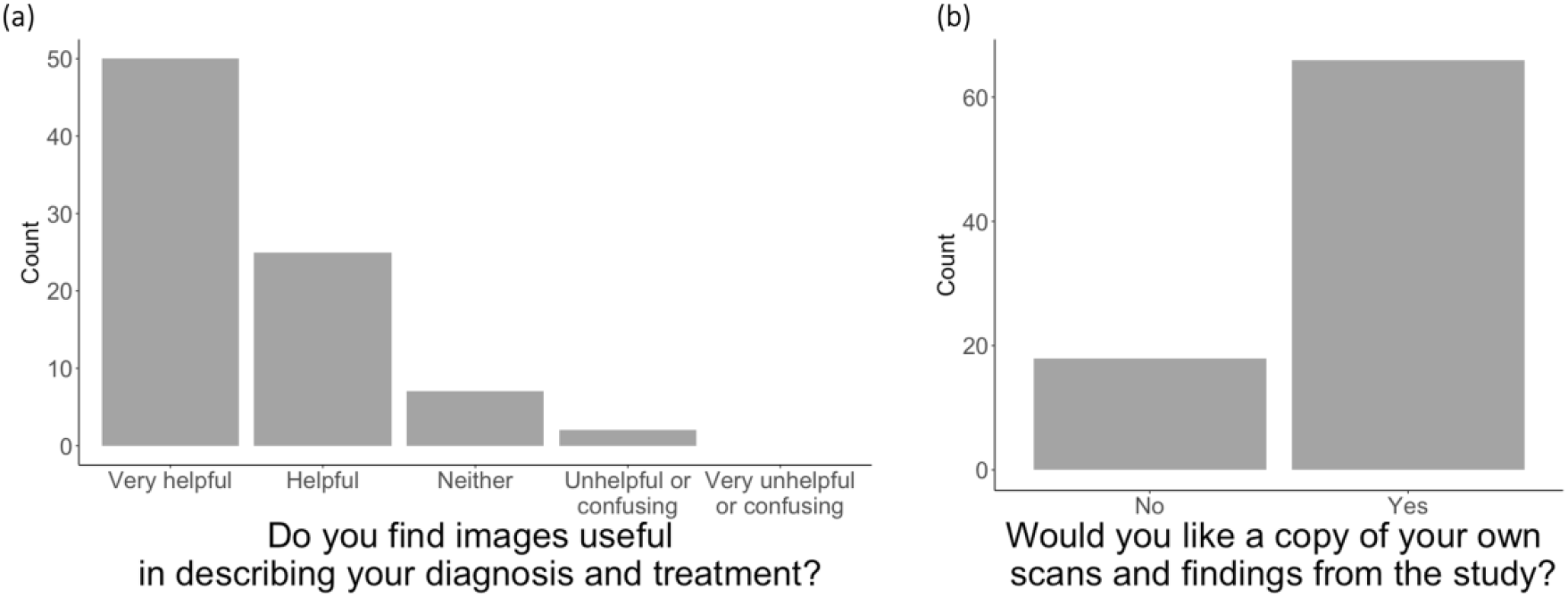
Patient reported understanding of diagnosis and treatment following provision of imaging report. 89% of participants found the additional images helpful or very helpful in describing their diagnosis and treatment. 78% of participants confirmed that they would like a copy of scans and findings from the study.

### Expected length of stay

Patients were asked about their expected length of stays following the treatment plan explanation with the support of the patient-friendly mpMRI report. The mean of the expected length of stay was 5.6 days, closely reflecting the mean actual length of stay of 5.3 days.

## Discussion

In this real-world evaluation of the impact of incorporating quantitative multi-parametric imaging as a marker of liver health for surgical planning we observed three key findings. First that the poor liver health was underestimated in 40% of patients about to undergo surgery; secondly that using mpMRI to assess liver health over and above the standard clinical work up, resulted in changes to the surgical plans in terms of both a more radical and more extensive procedure, personalising further the patient management; and third, that this impacts positively on the number of patients who experienced prolonged stay in hospital.

### Clinical Outcomes

The use of quantitative multiparametric MR imaging (mpMRI) to evaluate the health of the liver prior to surgical decision making resulted in only one patient requiring an extended (>14 days) stay in hospital, a dramatic reduction from a standard of care comparator dataset where 5% of patients required hospital care for over 14 days. This improvement in patient outcomes is a result of the additional clinical information indicating the health of the liver, reported with spatial information. Patients who were shown with mpMRI to have a liver with a high level of fat or liver disease activity underwent a more conservative resection or a two-stage resection; the perceived risk of poor post operative recovery enabled the selection of a safer treatment strategy. Only two patients had been previously diagnosed with parenchymal liver disease, while 23 patients were shown to have biomarkers of liver fat or liver disease activity using mpMRI.

Conversely, eight patients were revealed to have an unexpectedly healthy liver, as indicated by mpMRI showing low liver fat and cT1 within the normal range. This then supported the decision for a more extensive resection with more curative intent without the perceived risk of insufficient remnant liver. None of these six patients developed post-hepatectomy liver failure. Extended rounds of neoadjuvant chemotherapy ahead of an operation often raise concern, owing to the incidence of chemotherapy-associated steatohepatitis, which can be difficult to diagnose. mpMRI enables a clear evaluation of liver health such that a surgical strategy can be confidently planned. In four patients for whom an unexpectedly healthy liver was observed with mpMRI, their treatment was changed to a single extensive resection as opposed to a two stage treatment; this avoided the unnecessary second hospital visit of the two stage approach and the associated impacts on the patient and healthcare costs.

### Patient centred outcomes

mpMRI reports promote intuitive and fast understanding of disease activity of single segments and of the whole liver. Patient centred care including the treatment explanation supported by imaging reports resulted in a good understanding by patients of their disease and appropriate treatment steps. As a result, each patient’s expected length of stay reflected the actual length of stay. Several studies suggest an important influence of patient expectations on outcomes; it has been shown that the expectation of a shorter length of stay resulted in a faster convalescence of patients after major liver surgery^18^. In the future, referring to Hepatica based length of stay expectations rather than the national median length of stay, may foster quicker convalescence.

### Broader implications

Liver disease is a modifiable risk factor for patients undergoing complex liver surgery. The presence of significant parenchymal liver disease and low future liver remnant is associated with poor post-operative outcomes. As liver disease treatments including DVE, but also new pharmacological agents (e.g. tirzepatide, Eli Lilly) become available, we can begin to determine optimal pre-operative preparation for individual patients (personalised medicine).

Results shown here have broader consequences for the use of quantitative liver characterisation and the importance for controlling parenchymal liver disease at an early stage. The first stages of the spectrum of liver diseases have been shown to be reversible, including simple steatosis and non-alcoholic steatohepatitis, through dietary and pharmacological interventions. Chemotherapy-associated steatohepatitis in this group of patients is an acute condition particularly relevant in patients being considered for liver resection. Guidelines across various territories indicate that liver health should be evaluated prior to consideration for liver resection; however, the diagnostic accuracy and clinical usefulness of the recommended test (Child-Pugh Score) is best suited for end-stage liver disease. We have shown best-in-class diagnostic accuracy for early-stage liver disease across multiple disease aetiologies.

This therefore indicates a greater need for the role of Hepatologists within the care for patients with primary and secondary liver cancer. Accurate diagnosis and clinical control of parenchymal liver disease should be a part of all patients prior to assessment of eligibility for surgical resection. The medium-long term projections for the incidence of NAFLD-HCC are a cause for great concern and reiterate the as yet under-served clinical need for closer integration of hepatology, modern radiology and surgery within the multidisciplinary team management of patients being considered for liver resection.

Future work will be to examine long-term cancer-specific survival benefits of all patients who underwent more extensive resections. Limitations include data collected in a high-performing specialist centre which suggests the need to validate these results nation-wide. Future work will evaluate full healthcare resource utilisation and build an economic model to determine the level of cost saving that Hepatica creates.

## Supporting information

STROBE Checklist

## Data Availability

All data produced in the present study are available upon reasonable request to the authors

